# Diagnostic accuracy of CEUS in identifying Solid Pancreatic Lesions:- A meta analysis

**DOI:** 10.1101/2023.08.12.23294023

**Authors:** Dev Desai, Shimolee Patel, Hetvi Shah, Abhijay Shah, Anushka Verma, Maria Eleni Malafi

## Abstract

**Background:** Solid pancreatic lesions are crucial to identify because of their high incidence rate and their poor survival rate. Surgical biopsy, ultrasonography, computed tomography, MRI, and PET-CT are examples of diagnostic tools. Although common, endoscopic ultrasonography guided biopsy carries a risk of needle track seeding. A more effective and affordable method for determining the differential diagnosis of solid pancreatic lesions is contrast-enhanced ultrasonography (CEUS). CEUS is a less nephrotoxic method that uses a contrast chemical to distinguish between teratomas, benign tumors, and neuroendocrine tumors. The goal of this meta-analysis is to evaluate how well CEUS can identify solid pancreatic lesions for use in clinical diagnostic procedures.

**Methodology:** Medical literature comprehensively searched and reviewed without restrictions to particular study designs, or publication dates using PubMed, Cochrane Library and Google Scholar databases for all relevant literature. The extraction of necessary data proceeded after specific inclusion and exclusion criteria were applied. Meta Analysis included 27 RCTs and 3061 patients, and analyzed using the QualSyst yool. The risk of bias was evaluated by using QUADAS-2 analysis.

The statistical software packages MetaDiSc 1.4, RevMan (Review Manager, version 5.3), SPSS (Statistical Package for the Social Sciences, version 20) and Excel in Stata 14 were used to perform the statistical analyses.

**Result:** According to the findings of four studies, CEUS demonstrates high sensitivity, with values equal to or above 95%, and one study indicates specificity above 95%. True Positive (TP) and True Negative (TN) values are reported as 2080 and 621, respectively, while False Positive (FP) and False Negative (FN) values are noted as 124 and 236. With a 95% confidence interval, CEUS sensitivity is calculated as 0.90 (range: 0.89 to 0.91) and specificity as 0.83 (range: 0.80 to 0.86). The positive predictive value (PPV) of CEUS is estimated at approximately 94.3%. These results highlight CEUS as a promising tool for diagnosing pancreatic lesions.

**Conclusion:** The study concluded that CEUS (Contrast-Enhanced Ultrasound) is an important diagnostic test for pancreatic lesions. This is due to their high sensitivity and specificity, along with other aspects like enhanced visualization, real-time imaging, and safety benefits. Additionally, CEUS is cost-effective, making it a practical choice in healthcare settings with budget constraints.

Thus, CEUS remains a valuable asset for healthcare professionals in their efforts to accurately diagnose pancreatic lesions.

## INTRODUCTION

Solid pancreatic lesions include pancreatic ductal adenocarcinoma, neuroendocrine tumors, focal chronic pancreatitis, solid pseudopapillary tumor, pancreatic metastasis and many more, of which the most common is the pancreatic ductal adenocarcinoma. While looking at the ratings worldwide, it has an incidence rate of 13th among other malignant tumors. Till date it is considered one of the deadliest cancers, ranking fourth among the leading causes of death due to cancer in the United States. The 5-year survival rate for pancreatic cancer has been estimated to be around 6% owing to the limited availability of effective treatment options and drug therapy and poor prognosis with surgical resection.[1]

These tumors generally progress insidiously with minimal symptoms and tend to present in advanced stages when the condition worsens and resection is less practical. Thus, it becomes vital to diagnose the tumor as early as possible so that effective management and monitoring can be ensured.

Many diagnostic tools are being used to assess the tumor, ranging from surgical biopsy to various imaging tools like ultrasound (US), computed tomography (CT), magnetic resonance imaging (MRI) and positron emission tomography-computed tomography (PET-CT).[1], [2] Endoscopic ultrasound guided biopsy has been gaining popularity for the diagnosis of pancreatic lesion but there is a risk of needle track seeding which is concerning.[3]. While biopsy is considered the gold standard for definite diagnosis, non-invasive imaging methods play a pivotal role in diagnosis and evaluation of these tumor. Of the various imaging modalities, CT scan is the most widely used clinically. But while these methods come with their advantages, there are many downsides to them too, one of it being the high cost of getting these imaging done.

A relatively cheaper and superior option is the Contrast-enhanced ultrasound (CEUS), [1]especially for the differential diagnosis of solid pancreatic lesions. CEUS uses a contrast agent consisting of microbubbles of air or other gases, which is injected intravenously into the circulation to get real time imaging of the large vessels as well as the microvasculature. It has the advantage of being able to differentiate the origins of the lesions and evaluate their vascularity in real-time. Neuroendocrine tumor, benign tumors and teratoma are seen as hyperenhancement or isoenhancement while malignant tumors like the ductal adenocarcinoma are seen as hypo-enhancement[4] Further, CEUS has the advantage of being less nephrotoxic as the contrast agent used for CEUS is excreted via lungs instead of the kidneys. Taking all these advantages into account, this meta-analysis aims to ascertain the accuracy of CEUS to differentiate among various solid pancreatic lesions for its clinical application as a diagnostic tool.

### PRISMA Flowchart

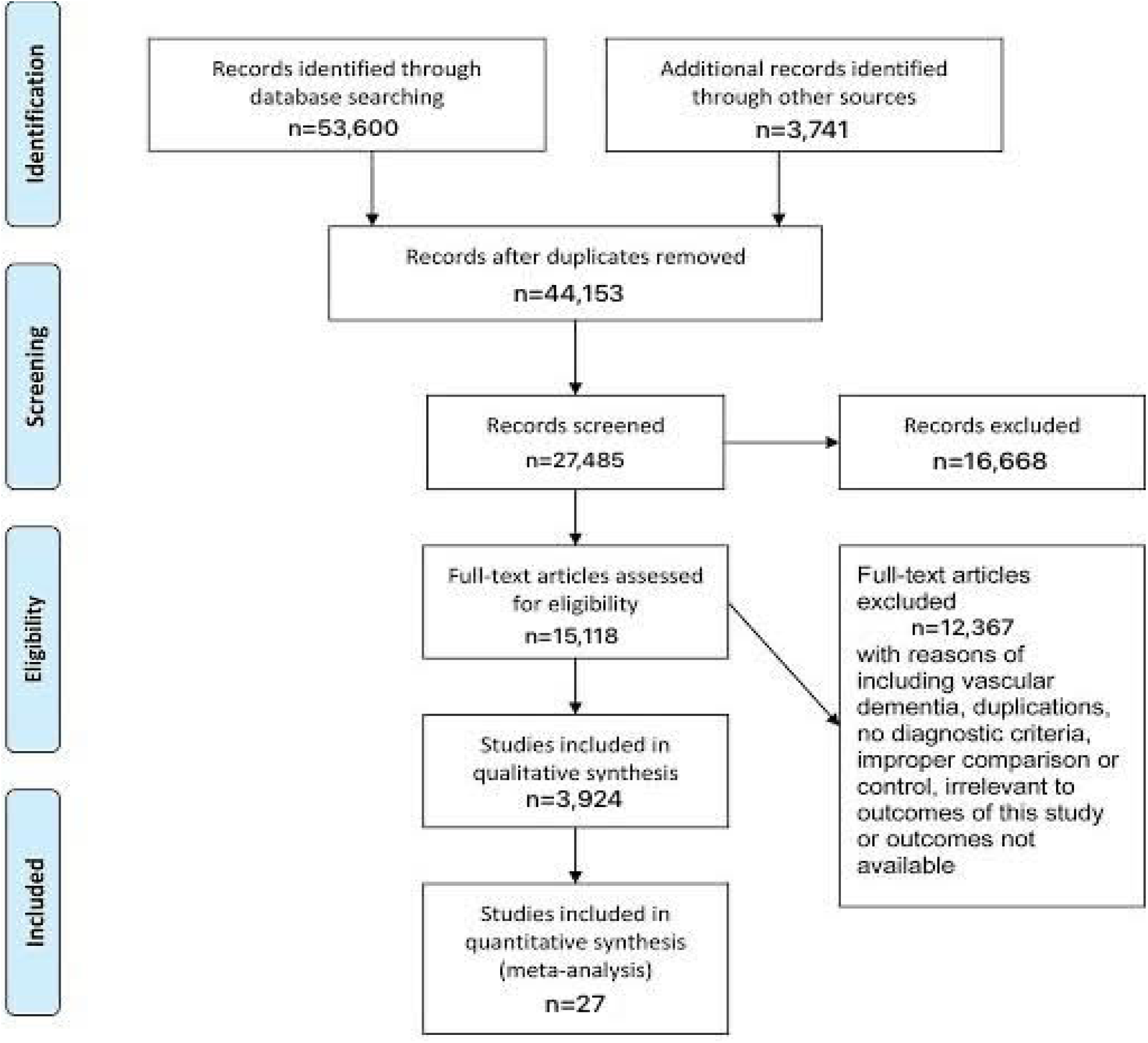

## METHODOLOGY

### DATA COLLECTION

For the collection of the data, a search was done by two individuals using PubMed, Google Scholar, and Cochrane Library databases for all relevant literature. Full - Text Articles written only in English were considered.

The medical subject headings (MeSH) and keywords ‘Contrast Enhanced Ultrasound’, ‘CEUS’, and ‘Pancreatic Lesions’ were used. References, reviews, and meta-analyses were scanned for additional articles.

### INCLUSION AND EXCLUSION CRITERIA

Titles and abstracts were screened, and duplicates and citations were removed. References of relevant papers were reviewed for possible additional articles. Papers with detailed patient information and statically supported results were selected.

We searched for papers that show more accurate diagnoses, where procedure being considered was CEUS. The inclusion criteria were as follows: (1) studies that provided information about the accurate diagnosis of solid pancreatic lesions with CEUS while main reference standard was biopsy or surgical pathology; (2) studies published in English; (3) Studies wherein true positive (TP), false positive (FP), false negative (FN), and true negative (TN) rates were obtained or calculated to construct the 2□×□2 contingency table. The exclusion criteria were: (1) articles that were not full text, (2) unpublished articles, and (3) articles in other languages.

### DATA EXTRACTION

Each qualifying paper was independently evaluated by two reviewers. Each article was analyzed for the number of patients, their age, procedure modality, and incidence of the pre decided complications. Further discussion or consultation with the author and a third party was used to resolve conflicts. The study’s quality was assessed using the modified Jadad score. In the end, According to PRISMA, a total of 27 RCTs with a total of 3061 patients were selected for further analysis.

### ASSESSMENT OF STUDY QUALITY

Using the QualSyst tool, two writers independently assessed the caliber of each included study. This test consists of 10 questions, each with a score between 0 and 2, with 20 being the maximum possible overall score. Two authors rated each article independently based on the above criteria. The interobserver agreement for study selection was determined using the weighted Cohen’s kappa (K) coefficient. For deciding the bias risk for RCTs, we also employed the Cochrane tool. No assumptions were made about any missing or unclear information. there was no funding involved in collecting or reviewing data. Heterogeneity and the diagnostic accuracy of CEUS were performed. the latter of which was calculated by pooled estimates of sensitivity, specificity, and diagnostic odds ratio (DOR) with corresponding 95% confidence intervals (CIs). Heterogeneity was detected by Cochrane Q test and I2 statistics, with P□<□0.1 or I2□>□50%, indicate a significance in heterogeneity. Furthermore, if there was significant heterogeneity (I2□>□50% or P□≤□.05), the random-effects model (DerSimonian-Laird method) was preferred over the fixed-effects model (Mantel-Haenszel method); otherwise, the fixed-effects model was the first choice.

### STATISTICAL ANALYSIS

The statistical software packages RevMan (Review Manager, version 5.3), SPSS (Statistical Package for the Social Sciences, version 20), Google Sheets, and Excel in Stata 14 were used to perform the statistical analyses. The data was obtained and entered into analytic software. Fixed-or random-effects models were used to estimate Sensitivity, Specificity, positive predictive value (PPV), diagnostic odds ratios (DOR), and relative risk (RR) with 95 percent confidence intervals to examine critical clinical outcomes (CIs). Diagnosis Accuracy and Younden Index were calculated for each result. Individual study sensitivity and specificity were plotted on Forest plots and in the receiver operating characteristic (ROC) curve. The forest plot and Fagan’s Nomogram were used to illustrate the sensitivity and specificity of different papers.[5]

### BIAS STUDY

The risk of bias was evaluated by using QUADAS-2 analysis. This tool includes 4 domains as Patient selection, Index test, Reference standard, Flow of the patients, and Timing of the Index tests. The summary of publication bias is shown in the following charts. The publication bias in patient selection was low in 17 high in 1 and unclear in 9. The index test was low in 25 and high in 1 paper. While the reference standard was low in 14, high in 1, and unclear in 12. The flow and timing were low in 20 and unclear in 7. The applicability concerns in patient selection were low in 10, high in 6, and unclear in 11. Reference standard was low in 11, high in 4, and unclear in 12 respectively. The index test was low in 23, unclear in 3 and high in 1 paper.

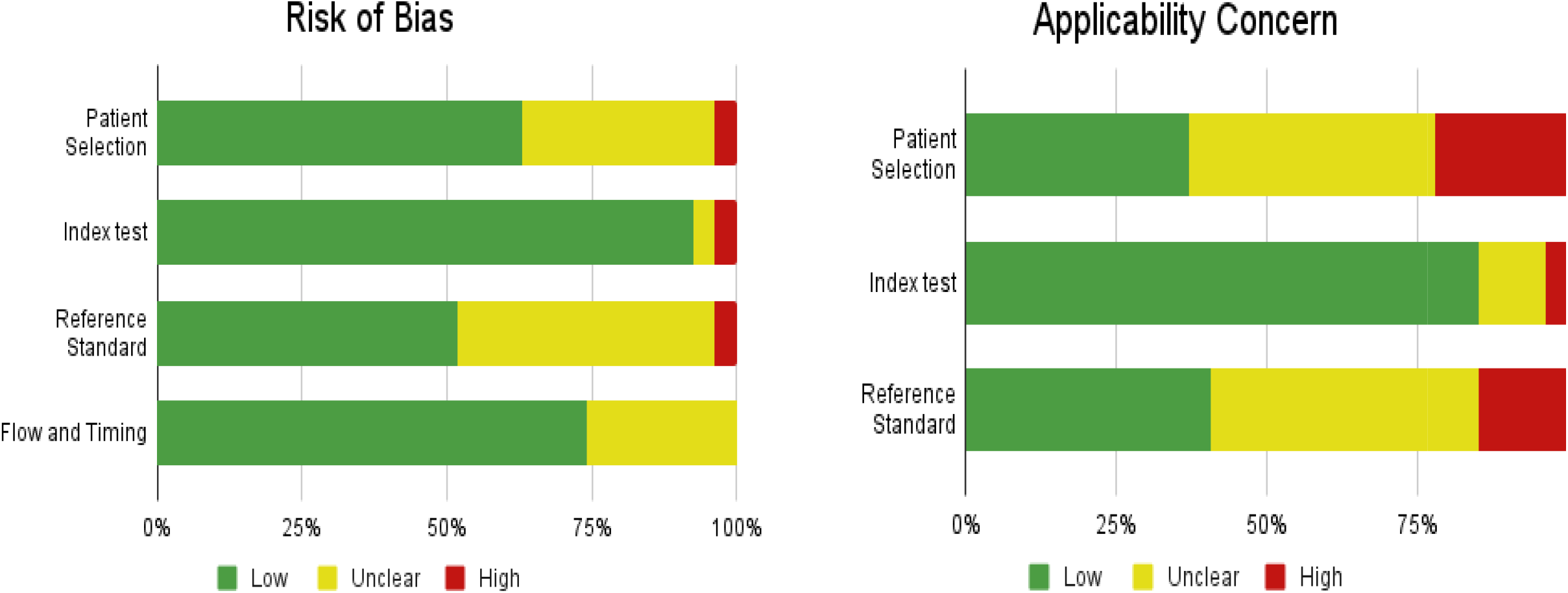

## RESULT

Here, Table 1 describes all the descriptions of papers used for the study regarding the use of CEUS in the diagnosis of Solid Pancreatic Lesions. In Figure 2, the Pooled Sensitivity Values for all papers being considered can be observed and compared amongst each other, while Figure 3 serves the same purpose in the context of Pooled Specificity Values. Figure 4 denotes the pooled Diagnostic Odds Ratio for the application of CEUS. The same is illustrated in the SROC curve. (Figure 5). A total of 27 RCTs with 3061 subjects were selected for the study, out of which 4 studies showed sensitivity above or equal to 95%, and 1 study showed specificity above 95%. The value of True Positive (TP) was 2080, that of True Negative (TN) was 621, that of False Positive (FP) was 124, and that of False Negative (FN) was 236. With a confidence interval 95%, sensitivity, specificity and positive predictive values were calculated. The sensitivity of CEUS is 0.90, with a CI of 95% in a range of 0.89 to 0.91. The specificity of CEUS is 0.83, with a CI of 95% in a range of 0.80 to 0.86.

**Table 1.**
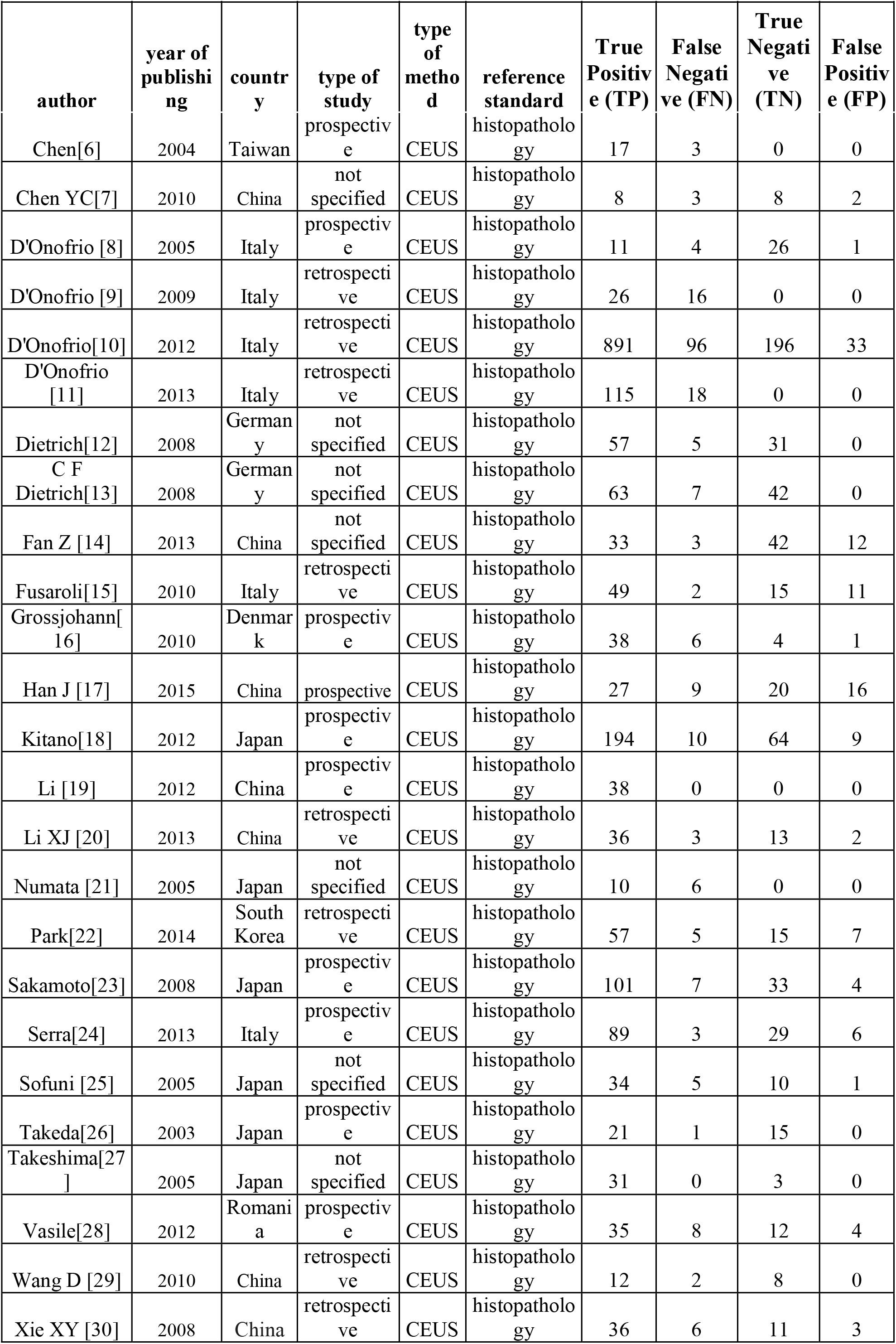
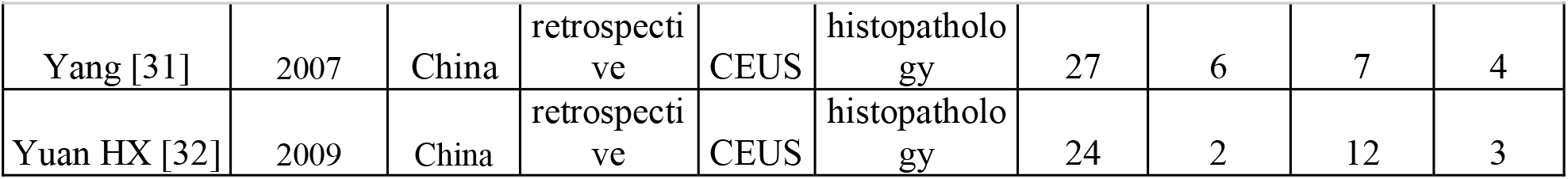

**Figure 2:**
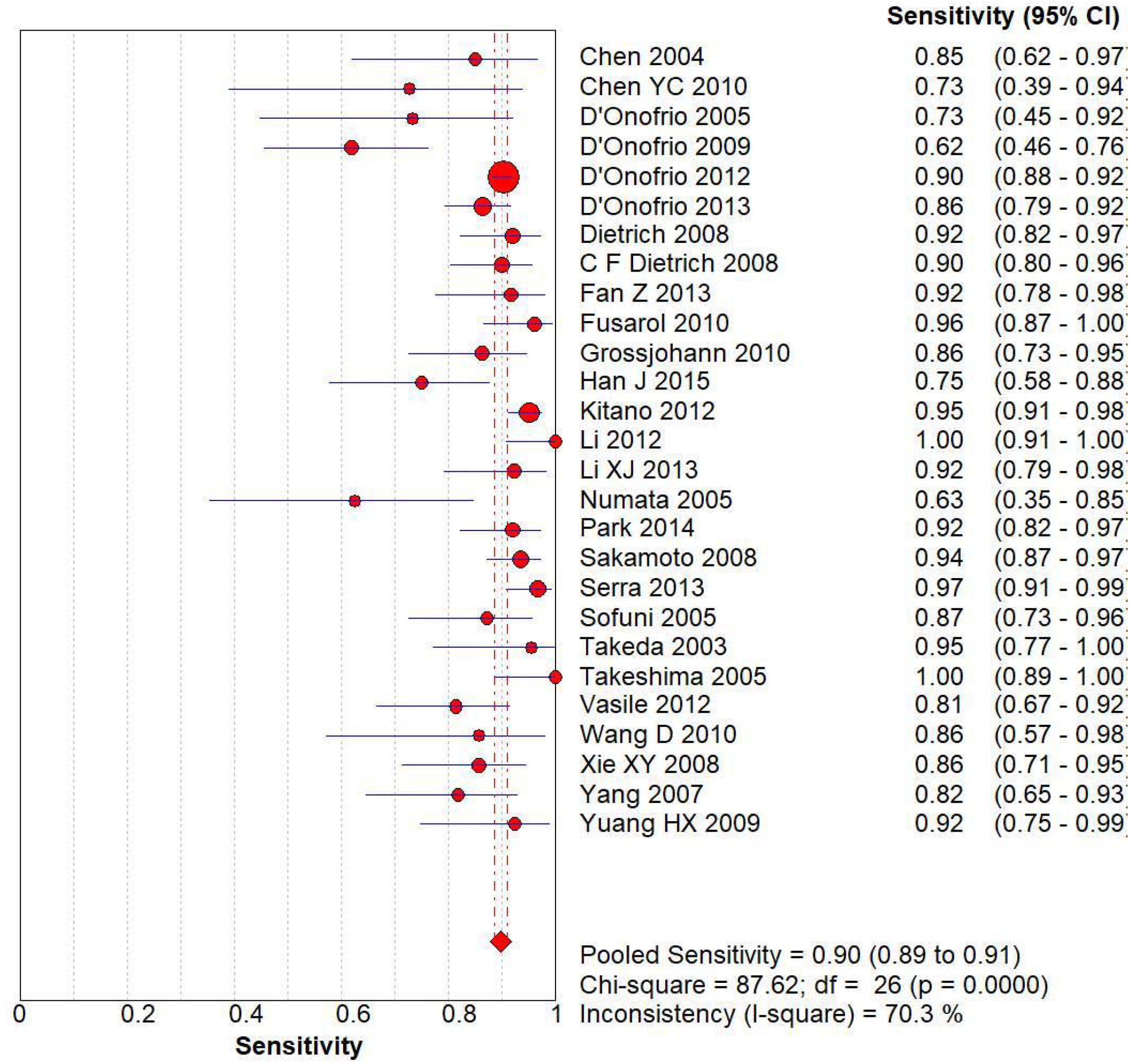
The forest chart summary for pooled sensitivity values.

**Figure 3:**
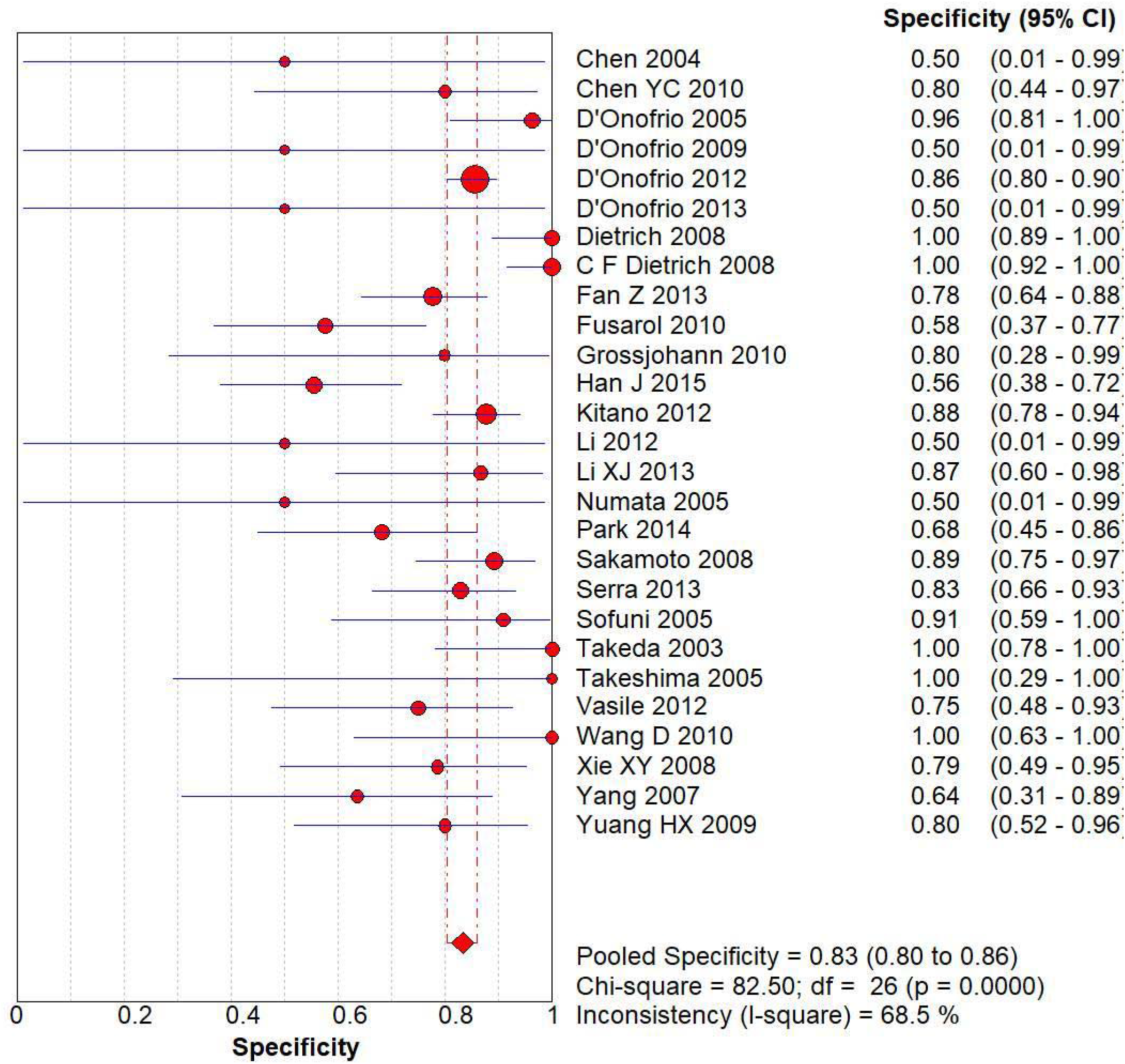
The forest chart summary for pooled specificity values.

**Figure 4:**
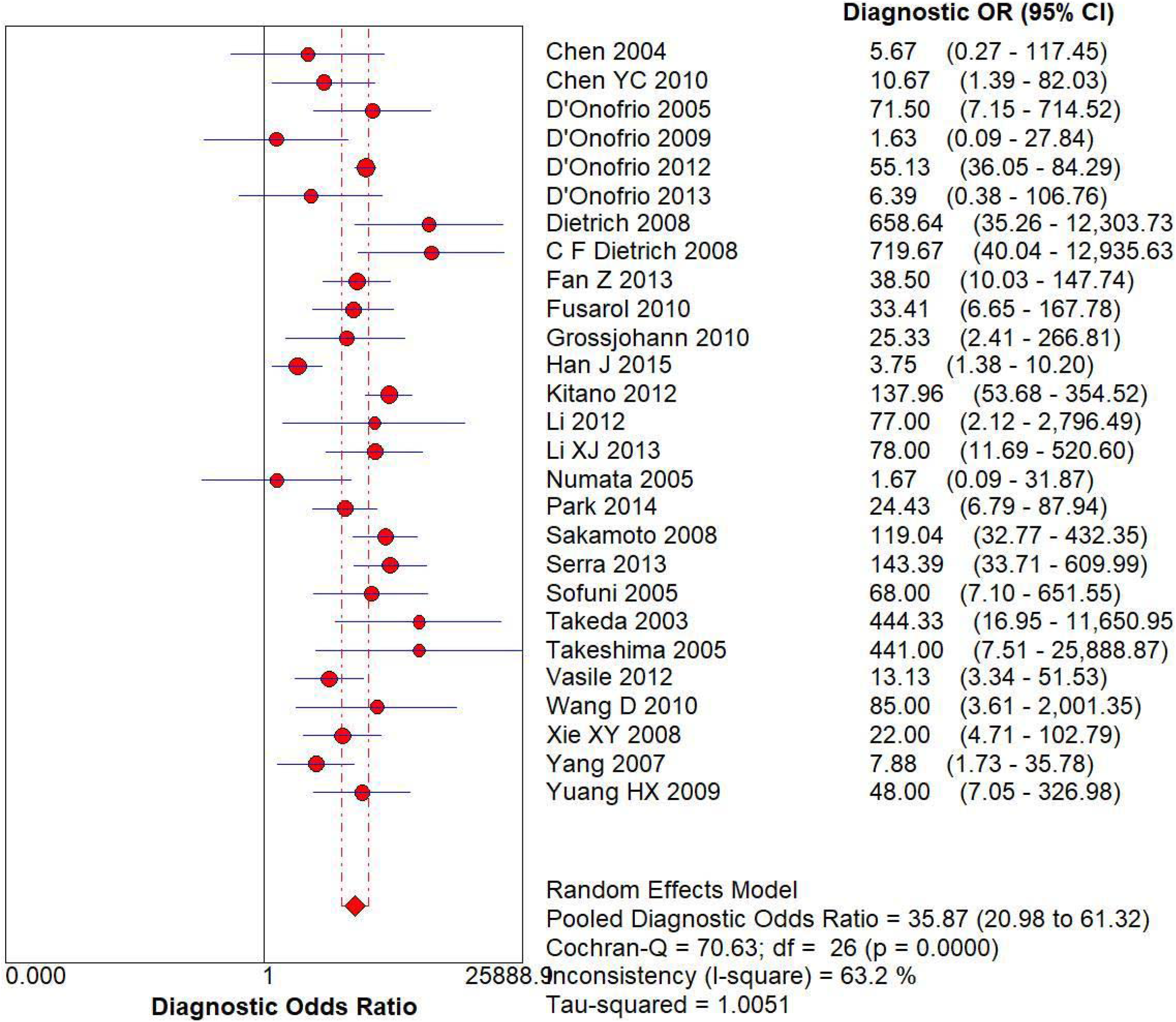
The forest chart summary for pooled Diagnostic Odds Ratio.

**Figure 5:**
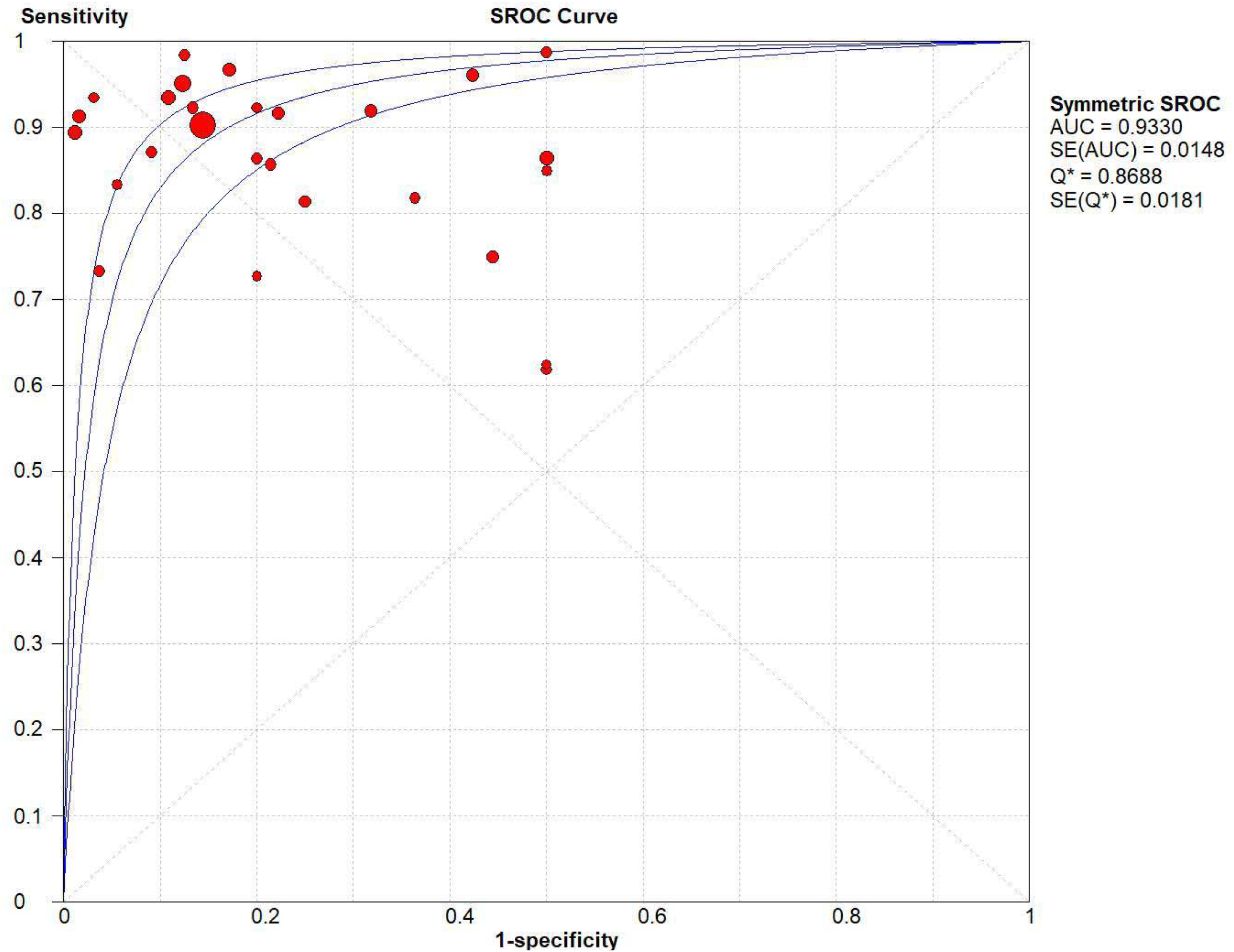
The SROC plot summary for CEUS for Solid Pancreatic Lesions.

The Cochrane Q was calculated with p-value<0.0001 and I^2^ = 63.2% for the Diagnostic Odds ratio and hence Random Effect model was used.

Figure 5 shows the summary of the ROC curve. It shows that the area under the curve for CEUS was 0.9390 and the overall diagnostic odds ratio (DOR) was 35.87 with the diagnostic accuracy and Younden Index being 0.884 and 0.736, respectively. For the SROC curve, the Index Q* intersection was at 0.8688 with Standard Error of 0.0181

Figure 6 describes the summary of Fagan plot analysis for all the studies considered for CEUS, showing a prior probability of 76% (3.1); a Positive Likelihood Ratio of 5.4; a probability of post-test 94% (16.8)); a Negative likelihood ratio of 0.12, and a probability of post-test 27% (0.4).

**Figure 6:**
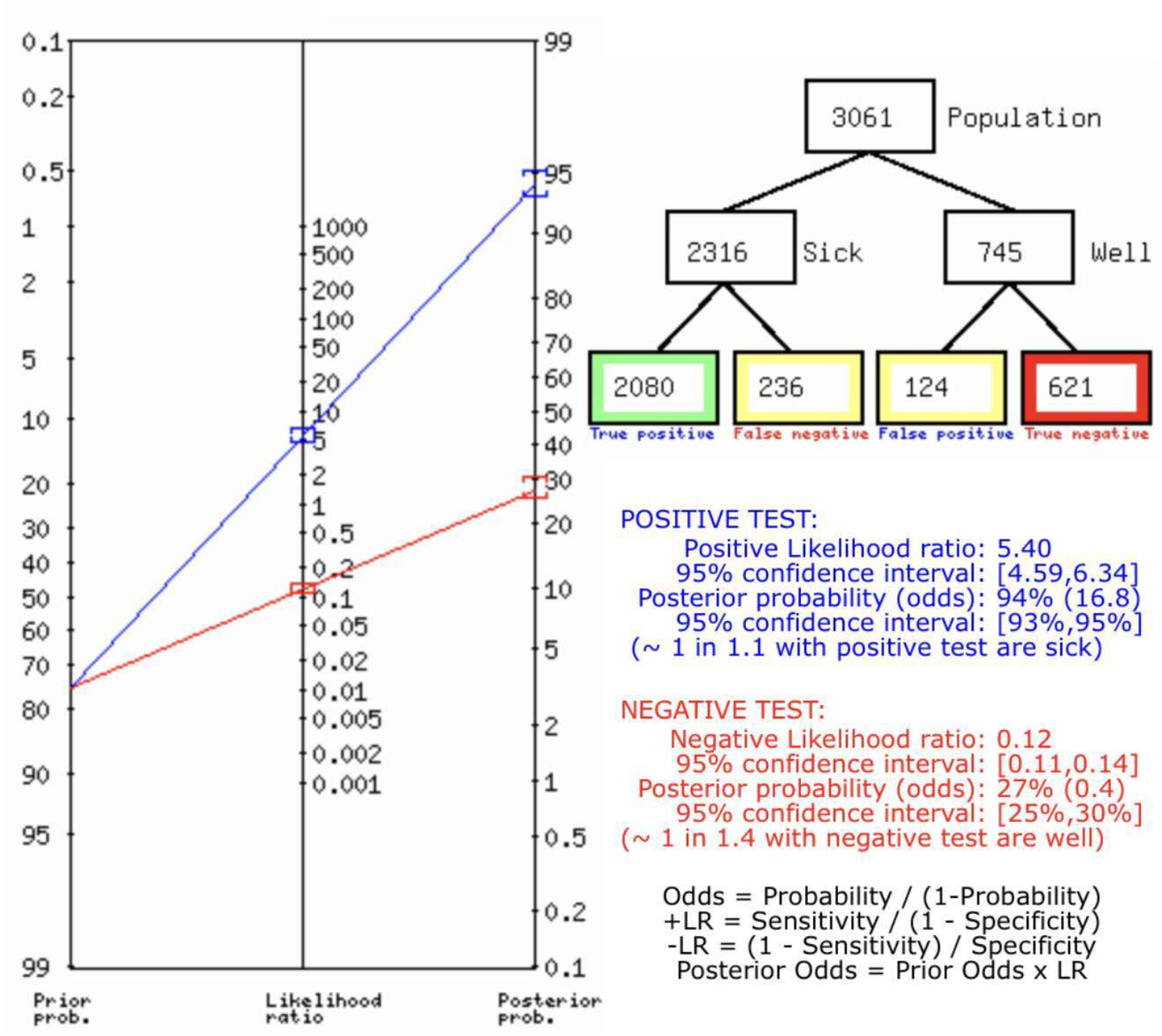
Fagan’s Analysis for CEUS for Solid Pancreatic Lesions.

**Figure 7:**
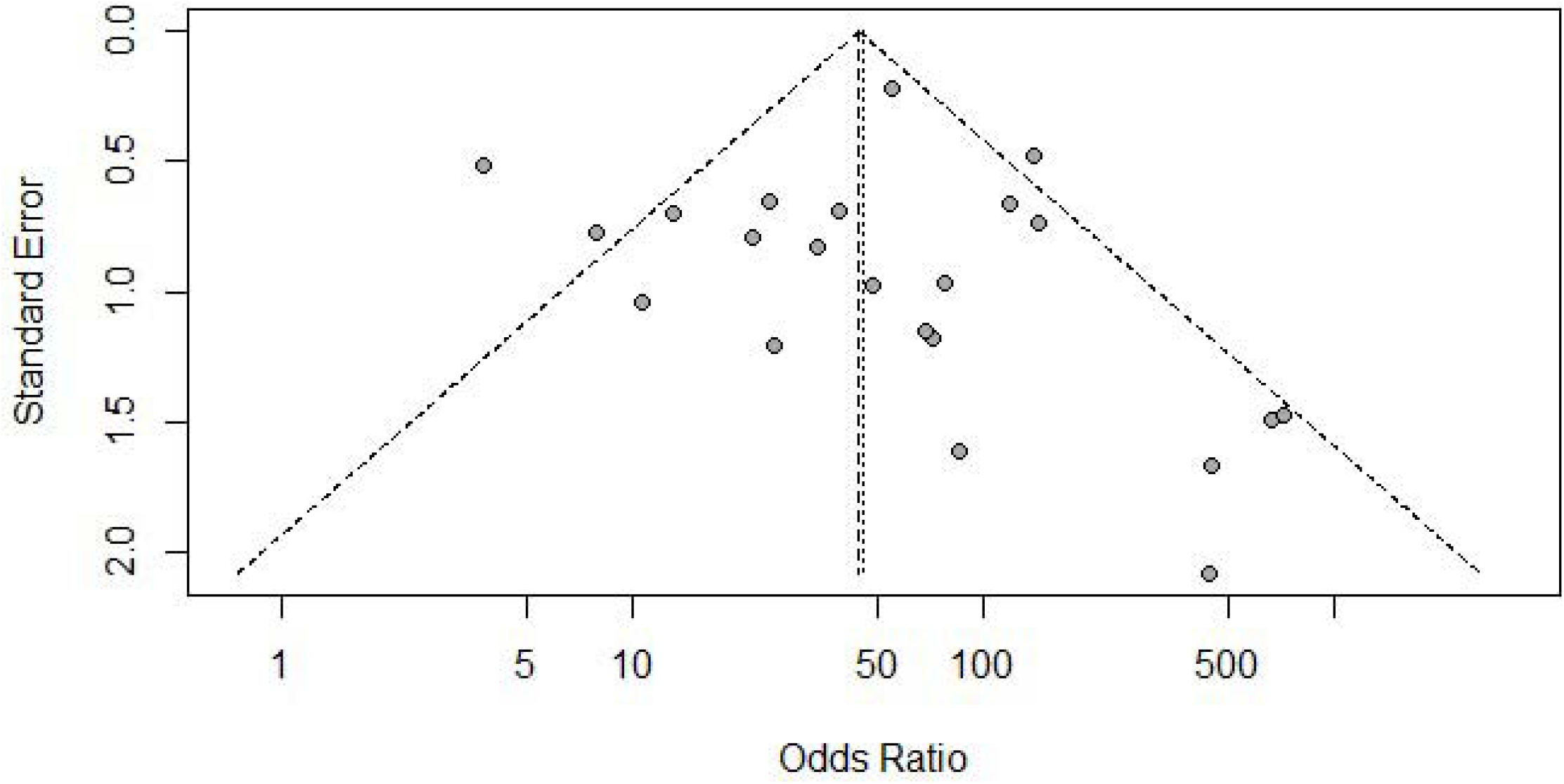
Symmetrical Funnel plot for Publication bias.

## DISCUSSION

In this meta-analysis our primary goal is to emphasis on the diagnostic accuracy of CEUS to identify a solid pancreatic lesion. After taking into consideration the results of 3061 subjects by intensive selection of the appropriate research papers, it has been found that CEUS has a pooled sensitivity of 90%, which means that there is 90% chance that the patient will be correctly diagnosed with the specific type of pancreatic lesion using CEUS, while 10% chance of the diagnosis being incorrect. On the other hand, CEUS has a pooled specificity of 83% which shows that every 83 people out of 100 will have a negative result on CEUS and 17 people will be wrongly diagnosed with a positive result.

Out of all the solid pancreatic lesions, the most common is the pancreatic adenocarcinoma which shows up as hypoenhancement on CEUS. Currently the most widely used imaging method for diagnosis of the pancreatic adenocarcinoma is the CT scan, which has a sensitivity of 81% and specificity of 43%. MRI also has comparable accuracy for diagnosing pancreatic adenocarcinoma with sensitivity of 89% and specificity of 63%, but it has the drawback of being a costly diagnostic tool which has limited availability in clinical centers. The endoscopic ultrasound guided fine needle aspiration (EUS-FNA) is a superior method for diagnosing the pancreatic adenocarcinoma and obtaining material for analysis with a sensitivity and specificity of 85% and 98% respectively. But it is an invasive method with risk of bleeding and needle track seeding of the pancreatic tumor cells. [33]Hence, CEUS proves to have a good diagnostic yield for evaluating pancreatic adenocarcinoma along with the advantage of being a non-invasive method with less associated pain and relatively low cost compared to other diagnostic methods.

CEUS provides a dynamic contrast with the use of a contrast agent consisting of microbubbles of air or other gases, which is injected intravenously into the circulation. It has an arterial phase of around 45 seconds followed by a venous phase of about 3 minutes. Unlike CECT and CEMR, there is no equilibrium phase in CEUS as it uses a purely intravascular contrast agent. The detection of vascularity in CEUS is done in the arterial phase with benign lesions and neuroendocrine tumors showing up as hyperenhancements or isoenhancement and malignant lesions like pancreatic adenocarcinoma presenting as hypoenhancement. [4] This variation in vascularity is used for the differential diagnosis of the solid pancreatic lesions with hypovascularity being a sign for malignancy.

With the advent of newer ultrasound contrast agents which overcomes the limitations of early breakdown of the microbubble that was seen in the previous contrast agents used for ultrasound, it has become possible to evaluate the lesions in real time using contrast-enhanced ultrasonography. [1] CEUS is an accessible method which is also comfortable to the patients. It does not cause any harmful effects on the kidney or thyroid as the contrast agent used for CEUS is excreted via lungs and does not contain iodine unlike seen in the other contrast based diagnostic methods like contrast-enhanced computed tomography (CECT) or contrast enhanced magnetic resonance imaging (CEMR). There is no risk of radiation toxicity in CEUS either. [4] Although it is a relatively safer method, there is still risk of hypersensitivity reaction due to the contrast agent.

Based on the results obtained and the advantages discussed, we are confident in the ability of CEUS in diagnosing a solid pancreatic lesion, especially in patients who prefer a non-invasive method.

## Conclusion

In summary, CEUS (Contrast-Enhanced Ultrasound) presents itself as a highly valuable and effective diagnostic method. CEUS assists in characterizing lesions, guiding biopsies accurately, and monitoring treatment responses. It complements CT and MRI in providing valuable insights for precise diagnosis, tailored interventions, and improved patient outcomes.

The comprehensive analysis of 27 randomized controlled trials involving a total of 3061 subjects provides valuable insights into the diagnostic performance of Contrast-Enhanced Ultrasound (CEUS) in detecting pancreatic lesions. The research emphasizes that CEUS has the capability to serve as a precise and effective imaging technique for this intended use.

The calculated sensitivity and specificity values, supported by robust 95% confidence intervals, further validate CEUS’s diagnostic prowess in pancreatic lesion detection. The sensitivity of 0.90, within the range of 0.89 to 0.91, underscores its effectiveness in identifying true positive cases. Similarly, the specificity of 0.83, falling between 0.80 and 0.86, solidifies its ability to correctly identify true negative cases.

These values collectively demonstrate CEUS’s potential to serve as a reliable diagnostic tool in clinical practice. By amalgamating data from a multitude of trials, the study establishes CEUS as a sensitive, specific, and accurate imaging modality. The findings offer valuable guidance to clinicians and researchers alike, encouraging the continued exploration and utilization of CEUS for enhanced diagnostic capabilities in pancreatic lesion assessment.

## Data Availability

All data produced in the present work are contained in the manuscript

## References

[1] L. Ran, W. Zhao, Y. Zhao, and H. Bu, “Value of contrast-enhanced ultrasound in differential diagnosis of solid lesions of pancreas (SLP),” Medicine (United States), vol. 96, no. 28. 2017. doi: 10.1097/MD.0000000000007463.

[2] A. Pulumati, A. Pulumati, B. S. Dwarakanath, A. Verma, and R. V. L. Papineni, “Technological advancements in cancer diagnostics: Improvements and limitations,” Cancer Reports, vol. 6, no. 2. 2023. doi: 10.1002/cnr2.1764.

[3] B. S. Jani et al., “Endoscopic ultrasound-guided fine-needle aspiration of pancreatic lesions: A systematic review of technical and procedural variables,” North American Journal of Medical Sciences, vol. 8, no. 1. 2016. doi: 10.4103/1947-2714.175185.

[4] Y. E. Chung and K. W. Kim, “Contrast-enhanced ultrasonography: Advance and current status in abdominal imaging,” Ultrasonography, vol. 34, no. 1, 2014, doi: 10.14366/usg.14034.

[5] D. Desai, “Dev’s Formulae for Diagnostic Test Accuracy Meta-analysis,” medRxiv, p. 2023.03.27.23287186, Jan. 2023, doi: 10.1101/2023.03.27.23287186.

[6] C. H. Chen, C. C. Yang, Y. H. Yeh, and M. H. Huang, “Contrast-Enhanced Power Doppler Sonography of Ductal Pancreatic Adenocarcinomas: Correlation with Digital Subtraction Angiography Findings,” Journal of Clinical Ultrasound, vol. 32, no. 4, 2004, doi: 10.1002/jcu.20018.

[7] Chen YC, “Diagnosis of Focal Lesions of Pancreases in Contrast-enhanced Ultrasound and Comparison with Contrast-enhanced CT. {Master Thesis},” China Medical University, Shenyang, Liaoning, China;, 2010.

[8] M. D’Onofrio et al., “Contrast-enhanced ultrasonography better identifies pancreatic tumor vascularization than helical CT,” Pancreatology, vol. 5, no. 4–5, 2005, doi: 10.1159/000086540.

[9] M. D’Onofrio et al., “Resectable Pancreatic Adenocarcinoma: Is the Enhancement Pattern at Contrast-Enhanced Ultrasonography a Pre-Operative Prognostic Factor?,” Ultrasound Med Biol, vol. 35, no. 12, 2009, doi: 10.1016/j.ultrasmedbio.2009.06.1100.

[10] M. D’Onofrio et al., “Pancreatic multicenter ultrasound study (PAMUS),” Eur J Radiol, vol. 81, no. 4, 2012, doi: 10.1016/j.ejrad.2011.01.053.

[11] M. D’Onofrio et al., “Comparison between CT and CEUS in the diagnosis of pancreatic adenocarcinoma,” Ultraschall in der Medizin, vol. 34, no. 4, 2013, doi: 10.1055/s-0032-1325324.

[12] C. F. Dietrich, A. Ignee, B. Braden, A. P. Barreiros, M. Ott, and M. Hocke, “Improved Differentiation of Pancreatic Tumors Using Contrast-Enhanced Endoscopic Ultrasound,” Clinical Gastroenterology and Hepatology, vol. 6, no. 5, 2008, doi: 10.1016/j.cgh.2008.02.030.

[13] C. F. Dietrich, B. Braden, M. Hocke, M. Ott, and A. Ignee, “Improved characterisation of solitary solid pancreatic tumours using contrast enhanced transabdominal ultrasound (Journal of Cancer Research and Clinical Oncology DOI: 10.1007/s00432-007-0326-6),” Journal of Cancer Research and Clinical Oncology, vol. 134, no. 6. 2008. doi: 10.1007/s00432-007-0335-5.

[14] Z. Fan et al., “Application of contrast-enhanced ultrasound in the diagnosis of solid pancreatic lesions -A comparison of conventional ultrasound and contrast-enhanced CT,” Eur J Radiol, vol. 82, no. 9, 2013, doi: 10.1016/j.ejrad.2013.04.016.

[15] P. Fusaroli, A. Spada, M. G. Mancino, and G. Caletti, “Contrast Harmonic Echo-Endoscopic Ultrasound Improves Accuracy in Diagnosis of Solid Pancreatic Masses,” Clinical Gastroenterology and Hepatology, vol. 8, no. 7, 2010, doi: 10.1016/j.cgh.2010.04.012.

[16] H. S. Grossjohann et al., “Usefulness of contrast-enhanced transabdominal ultrasound for tumor classification and tumor staging in the pancreatic head,” Scand J Gastroenterol, vol. 45, no. 7–8, 2010, doi: 10.3109/00365521003702718.

[17] Han J, “The application of contrast-enhanced ultrasound in the diagnosis of focal pancreatic lesions. (Doctor Thesis), ” Chinese Academy of Medical Sciences, Beijing, China;, 2015.

[18] M. Kitano et al., “Characterization of small solid tumors in the pancreas: The value of contrast-enhanced harmonic endoscopic ultrasonography,” American Journal of Gastroenterology, vol. 107, no. 2, 2012, doi: 10.1038/ajg.2011.354.

[19] S. Li, P. Huang, H. Xu, K. Xu, and X. Wu, “Comparison of double contrast-enhanced ultrasound and MDCT for assessing vascular involvement of pancreatic adenocarcinoma: Preliminary results correlated with surgical findings,” Ultraschall in der Medizin, vol. 33, no. 7, 2012, doi: 10.1055/s-0031-1299429.

[20] Z. R. Z. X. et al. Li XJ, “ Combination of ultrasonic imaging technology with multislice CT enhancement technology in the diagnosis of pancreatic cancer and the clinical value of pancreatic resection judgment. Value of contrast-enhanced ultrasound in differential diagnosis of solid lesions of pancreas (SLP).,” J Qiqihar Univ Med;;4:545–6, 2013.

[21] K. Numata et al., “Contrast-enhanced sonography of pancreatic carcinoma: Correlations with pathological findings,” J Gastroenterol, vol. 40, no. 6, 2005, doi: 10.1007/s00535-005-1598-8.

[22] J. S. Park, H. K. Kim, B. W. Bang, S. G. Kim, S. Jeong, and D. H. Lee, “Effectiveness of contrast-enhanced harmonic endoscopic ultrasound for the evaluation of solid pancreatic masses,” World J Gastroenterol, vol. 20, no. 2, 2014, doi: 10.3748/wjg.v20.i2.518.

[23] H. Sakamoto, M. Kitano, Y. Suetomi, K. Maekawa, Y. Takeyama, and M. Kudo, “Utility of Contrast-Enhanced Endoscopic Ultrasonography for Diagnosis of Small Pancreatic Carcinomas,” Ultrasound Med Biol, vol. 34, no. 4, 2008, doi: 10.1016/j.ultrasmedbio.2007.09.018.

[24] C. Serra et al., “Contrast-enhanced ultrasound in the differential diagnosis of exocrine versus neuroendocrine pancreatic tumors,” Pancreas, vol. 42, no. 5, 2013, doi: 10.1097/MPA.0b013e31827a7b01.

[25] A. Sofuni et al., “Differential diagnosis of pancreatic tumors using ultrasound contrast imaging,” J Gastroenterol, vol. 40, no. 5, 2005, doi: 10.1007/s00535-005-1578-z.

[26] K. Takeda et al., “Contrast-enhanced transabdominal ultrasonography in the diagnosis of pancreatic mass lesions,” Acta radiol, vol. 44, no. 1, 2003, doi: 10.1034/j.1600-0455.2003.00009.x.

[27] K. Takeshima et al., “Comparison of IV contrast-enhanced sonography and histopathology of pancreatic cancer,” American Journal of Roentgenology, vol. 185, no. 5, 2005, doi: 10.2214/AJR.04.1588.

[28] T. A. Vasile et al., “Contrast enhanced ultrasound and computer tomography diagnosis of solid and mixed pancreatic tumors -analysis of confounders,” Journal of Gastrointestinal and Liver Diseases, vol. 21, no. 3, 2012.

[29] T. S. G. J. et al. Wang D, “ Contrast-enhanced ultrasonographic appearance of focal lesions of pancreas and the correlation with microvascular density,” Chin J Med Imaging Technol 25:2069–72., 2009.

[30] X. Y. Xie et al., “Role of contrast-enhanced ultrasound in the differentiation of solid focal lesions of pancreas,” Acta Academiae Medicinae Sinicae, vol. 30, no. 1, 2008.

[31] W. Yang, M. H. Chen, K. Yan, W. Wu, Y. Dai, and H. Zhang, “Differential diagnosis of non-functional islet cell tumor and pancreatic carcinoma with sonography,” Eur J Radiol, vol. 62, no. 3, 2007, doi: 10.1016/j.ejrad.2007.02.042.

[32] D. H. L. L. et al. Yuan HX, “ Value of contrast-enhanced ultrasound in solid neoplasms of pancreas.,” Shanghai Med Imaging, vol. 19, pp. 102–104, 2010.

[33] M. I. et al. Costache, “ ‘Which is the Best Imaging Method in Pancreatic Adenocarcinoma Diagnosis and Staging -CT, MRI or EUS?.,’” Current health sciences journal, vol. 43,2, pp. 132–136, 2017.

